# Longitudinal analysis of SARS-CoV-2 vaccine breakthrough infections reveal limited infectious virus shedding and restricted tissue distribution

**DOI:** 10.1101/2021.08.30.21262701

**Authors:** Ruian Ke, Pamela P. Martinez, Rebecca L. Smith, Laura L. Gibson, Chad J. Achenbach, Sally McFall, Chao Qi, Joshua Jacob, Etienne Dembele, Camille Bundy, Lacy M. Simons, Egon A. Ozer, Judd F. Hultquist, Ramon Lorenzo-Redondo, Anita K. Opdycke, Claudia Hawkins, Robert L. Murphy, Agha Mirza, Madison Conte, Nicholas Gallagher, Chun Huai Luo, Junko Jarrett, Abigail Conte, Ruifeng Zhou, Mireille Farjo, Gloria Rendon, Christopher J. Fields, Leyi Wang, Richard Fredrickson, Melinda E. Baughman, Karen K. Chiu, Hannah Choi, Kevin R. Scardina, Alyssa N. Owens, John Broach, Bruce Barton, Peter Lazar, Matthew L. Robinson, Heba H. Mostafa, Yukari C. Manabe, Andrew Pekosz, David D. McManus, Christopher B. Brooke

**Author notes:** These authors contributed equally (Ruian Ke, Pamela P. Martinez).

## Abstract

The global effort to vaccinate people against SARS-CoV-2 in the midst of an ongoing pandemic has raised questions about the nature of vaccine breakthrough infections and the potential for vaccinated individuals to transmit the virus. These questions have become even more urgent as new variants of concern with enhanced transmissibility, such as Delta, continue to emerge. To shed light on how vaccine breakthrough infections compare with infections in immunologically naive individuals, we examined viral dynamics and infectious virus shedding through daily longitudinal sampling in a small cohort of adults infected with SARS-CoV-2 at varying stages of vaccination. The durations of both infectious virus shedding and symptoms were significantly reduced in vaccinated individuals compared with unvaccinated individuals. We also observed that breakthrough infections are associated with strong tissue compartmentalization and are only detectable in saliva in some cases. These data indicate that vaccination shortens the duration of time of high transmission potential, minimizes symptom duration, and may restrict tissue dissemination.

## MAIN TEXT

Licensed vaccines against SARS-CoV-2 have already established a clear record of success in reducing case numbers and disease severity in areas that have achieved high vaccination rates. While vaccination clearly limits susceptibility to the virus, breakthrough infections can occur, as no vaccines are capable of eliciting sterilizing immunity, particularly against viruses that infect mucosal tissues^1^. Characterizing the unique features of breakthrough infections is critical for evaluating the potential for vaccinated individuals to transmit virus and understanding how vaccine-induced immunity suppresses viral replication and mitigates disease severity.

A study performed in Israel estimated that vaccination with the BNT162b2 (Pfizer/BioNTech) mRNA vaccine substantially reduced the potential for transmission among household contacts^2^. Other recent data have suggested that vaccinated individuals may shed virus (as measured by RTqPCR) at similar levels to unvaccinated individuals, particularly when infected with the Delta variant^3–5^. Some interpretations of these preliminary studies have suggested that the risk for secondary transmission is similar for vaccinated and unvaccinated individuals. However, most studies to date base their conclusions on cross sectional sampling of viral genome loads (as measured by RTqPCR), which may not directly translate to infectiousness^6^. In a cohort of longitudinally sampled participants who screened positive for SARS-CoV-2, we previously showed that the relationship between viral genome load and infectious virus load can vary greatly across individuals and over time, making the use of cross-sectional RTqPCR data problematic for estimations of infectiousness^7^. Therefore, longitudinal comparisons of viral genome shedding and infectious virus shedding across tissue compartments between vaccinated and unvaccinated individuals are needed to more accurately assess the effects of vaccination on viral dynamics and transmission potential.

Here, we present the longitudinal dynamics of SARS-CoV-2 infection in 23 individuals infected at varying stages of vaccination (6 fully vaccinated, *i*.*e*. enrolled at least 14 days after second mRNA vaccine dose or first J&J vaccine dose; 6 partially vaccinated, *i*.*e*. not fully vaccinated but enrolled at least 14 days past first mRNA vaccine dose; and 11 newly vaccinated individuals that enrolled less than 14 days after first vaccine dose (either mRNA or J&J)), captured at two study sites through daily nasal swab and saliva collection, along with symptom reporting (**Fig 1A-C**). These individuals were primarily infected with B.1.1.7 (Alpha) and P.1 (Gamma) variants, as enrollment in this study concluded before the widespread circulation of Delta at the study sites (**Table 1**)

**Figure 1:**
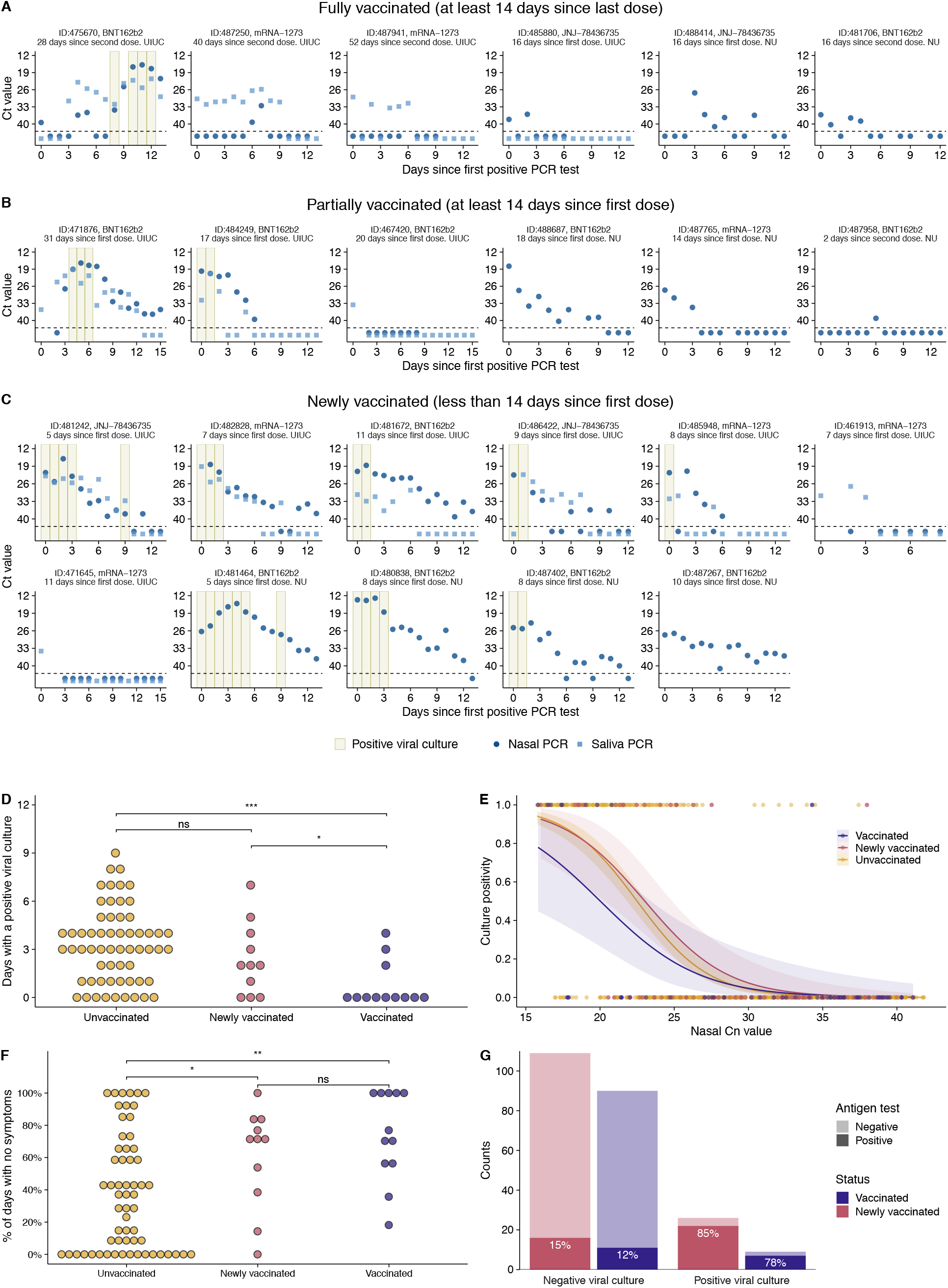
Viral dynamics in vaccinated individuals. **(A)** Temporal trends for the saliva RTqPCR (light blue squares), nasal swab RTqPCR (dark blue dots), and positive nasal swab viral culture results (tan bars) in fully vaccinated individuals that enrolled >/= 14 days after second mRNA vaccine dose or first J&J vaccine dose. The X axis shows days since the first positive PCR result. The Y axis indicates Ct values for saliva RTqPCR assay (covidSHIELD) and Cn values for nasal swab RTqPCR assay (Abbott Alinity). Horizontal dashed line indicates limit of detection of RTqPCR assays. For individuals at the NU study site, saliva samples were not collected thus only nasal swab data are shown. **(B)** Same data as in (A) but for partially vaccinated individuals that enrolled ≥14 days after first mRNA vaccine dose but were not yet fully vaccinated. **(C)** Same data as in (A) but for newly vaccinated individuals that enrolled < 14 days after first mRNA or J&J vaccine dose. **(D)** Numbers of days that vaccinated (combined fully and partially vaccinated individuals), newly vaccinated individuals, and unvaccinated individuals (from Ke et al.^7^) tested viral culture positive. ns, p>0.05; * p<0.05; ** p<0.01; *** p<0.001. **(E)** Association between the nasal Cn values and the probability of the sample being viral culture positive summarized across the vaccinated individuals, newly vaccinated individuals, and unvaccinated individuals (from Ke et al.^7^). Dots indicate individual viral culture results, 1 being a positive result, and 0 a negative result. The solid line and the shaded area are the mean and the confidence intervals, respectively, of a logistic regression fit. **(F)** Proportion of days post-enrollment (up to 14 days) that vaccinated, newly vaccinated, and unvaccinated individuals reported no symptoms. ns, p>0.05; * p<0.05; ** p<0.01; *** p<0.001. **(G)** Plot showing antigen FIA results from days where participants tested either positive or negative by viral culture. The text inside the bars indicates the percentage of antigen FIA results that were positive when concurrent viral culture sample was positive or negative.

5 out of 6 fully vaccinated individuals remained viral culture negative throughout their enrollment period, suggesting minimal shedding of infectious virus and little to no transmission risk. Moreover, the 5 individuals who remained viral culture negative had either undetectable or sporadic and low-level (generally Cn>35) viral genome loads in the nasal compartment.

Interestingly, in 2 (487941 and 487250) of the 3 viral culture negative individuals for which we collected both saliva and nasal samples, viral RNA was detectable in saliva for 5 to 10 days while remaining either undetectable (487941) or detectable at very low level for 2 days (487250) in nasal swabs. These data suggest that in 2 of the 4 fully vaccinated individuals for which both saliva and nasal swabs were collected, infection was initially established within the oral cavity or other saliva-exposed tissue site and was restricted from disseminating to the nasal passages. We did not observe a similar restriction of virus to saliva across 60 non-vaccinated individuals that we examined in a previous report^7^, suggesting that severe compartmentalization and tissue-restriction of virus may be a unique feature of vaccine breakthrough infections.

The one fully vaccinated individual (475670) that did test viral culture positive exhibited highly discordant patterns of viral shedding between saliva and nasal swabs. Viral genome loads expanded and declined in saliva samples over the first week of sample collection while remaining very low or undetectable in nasal swabs. At day nine post-enrollment, viral genome loads suddenly spiked in nasal samples and the individual began testing viral culture positive. This pattern is consistent with initial containment of the virus in saliva-associated tissue, followed by eventual viral breakthrough and dissemination to the nasal compartment.

Patterns of viral shedding in partially vaccinated individuals were more variable. Of the 6 individuals that were not considered fully vaccinated but enrolled *≥*14 days after receiving the first dose of mRNA vaccine, 2 only tested positive by RT-qPCR in a single sample (out of 13 or 15 total samples) (**Fig. 1B**), suggesting highly restricted infection with minimal transmission risk. In the other 3 individuals, viral shedding dynamics were indistinguishable from what we previously observed in unvaccinated individuals^7^, and 2 of these 3 tested viral culture positive on at least one day (**Fig. 1B**). Of the 11 individuals that enrolled within 14 days of receiving their first vaccine dose (“newly vaccinated”), most appeared similar to unvaccinated individuals with the exception of three that appeared to exhibit restricted shedding (**Fig. 1C**). These data are consistent with individual variation in the onset and magnitude of vaccine-mediated protection.

We directly compared duration of infectious virus shedding between fully and partially vaccinated individuals (combined here as “vaccinated” due to low numbers), newly vaccinated individuals, and unvaccinated individuals from our previous study (**Fig. 1D**)^7^. The total numbers of days that vaccinated individuals tested viral culture positive was significantly fewer than both newly vaccinated and unvaccinated groups, indicating that vaccination significantly reduces infectious virus shedding.

We also examined whether the relationship between nasal swab Cn value and viral culture status differed in vaccinated (both fully and partially), newly vaccinated, and unvaccinated individuals (from Ke et al^7^) (**Fig. 1E**). For samples with Cn values below 27, we found that the probability of being viral culture positive was lower for samples coming from vaccinated individuals versus newly vaccinated and unvaccinated individuals. These data suggest that for a given viral genome load (as measured by RTqPCR), vaccinated individuals may be less infectious than unvaccinated individuals, consistent with a recent report examining Delta breakthrough infections^8^. However, we must emphasize that this difference is not statistically significant, potentially due to both the relatively small number of samples from vaccinees and the fact that only 6 out of 12 individuals included in the vaccinated group were fully vaccinated at the time of enrollment. Regardless, these data further illustrate that Ct/Cn values cannot be used as a simple surrogate for infectious potential.

We next examined whether there were any differences in self-reported symptoms between vaccinated and unvaccinated individuals (using the 60 unvaccinated individuals previously reported^7^) (**Fig. 1F**). A Poisson regression shows that those who received at least one vaccine dose had significantly more days with no reported symptoms than the unvaccinated (p<0.0001). The mean proportion of study days with no symptoms was 0.74 in the vaccinated group dose compared with 0.37 in the unvaccinated group (range: 0 to 1 for both groups).

Finally, we examined the relationship between viral culture and antigen FIA results in vaccinated (fully plus partially) and newly vaccinated individuals **(Fig 1G)**. We observed that vaccinated and newly vaccinated participants tested positive by antigen FIA on 78% and 85% of the days on which they also tested positive by viral culture, suggesting that antigen FIA can be used to identify vaccine breakthrough infections with high transmission risk, especially if used as part of a serial screening program^9^. These results are consistent with our previous results in unvaccinated individuals as well as earlier cross-sectional studies examining the relationship between antigen tests positivity and infectious virus shedding^7,10,11^.

This study has several limitations that must be considered. First, the study cohort size is small, thus making it hard to draw firm quantitative conclusions. Second, our study cohort is biased towards breakthrough infections that were detected in our on-campus screening programs (saliva-based RTqPCR at UIUC, nasal swab-based LAMP assay at NU). Finally, enrollment in this study concluded before the arrival of the Delta variant at either study site. It remains unclear how well the effects of vaccination on viral infection dynamics that we describe apply to Delta variant breakthrough infections, given the unique features^12^ and enhanced transmissibility^13^ of this variant relative to the viruses we captured here.

Overall, our data suggest that vaccinated individuals are less likely to be shedding infectious virus at a given viral genome load and shed for a shorter period of time compared to unvaccinated and report fewer days of symptoms. We also show that some breakthrough infections in fully vaccinated individuals may be tissue restricted and only detectable through saliva screening. The clinical implications of compartmentalization are that testing (RT-PCR or antigen) based on nasal swabs may underestimate the true number of breakthrough infections, and that an important role of vaccine-elicited immunity may be restricting viral dissemination and thus limiting symptom severity and transmission potential. These data also further support a role for the oral cavity or other saliva-associated tissue sites as an initial site for SARS-CoV-2 infection prior to dissemination and replication of the virus in nasal passages in some individuals. Altogether, this study provides a set of high resolution data that ratify the role of the current SARS-CoV-2 vaccines not only in reducing severity of the disease, but also the infectiousness of individuals with breakthrough infections.

## METHODS

This study was approved by the Western Institutional Review Board, and all participants provided informed consent.

### Participants

University of Illinois at Urbana-Champaign (UIUC) enrollment site: All on-campus students and employees of the University of Illinois at Urbana-Champaign were required to submit saliva for RTqPCR testing every 2-4 days as part of the SHIELD campus surveillance testing program^14^. Those testing positive were instructed to isolate and were eligible to enroll in this study for a period of 24 hours following receipt of their positive test result. Close contacts of individuals who test positive (particularly those co-housed with them) were instructed to quarantine and were eligible to enroll for up to 5 days after their last known exposure to an infected individual. All participants were also required to have received a negative saliva RTqPCR result 7 days prior to enrollment.

Northwestern University (NU) enrollment site: All NU on-campus students were required to have nasal swab samples collected for LAMP testing once per week as part of the campus surveillance program. Those testing positive were required to go in the Health Service Quarantine and Isolation (QI) program for isolation. They were eligible for enrollment in this study within 24 hours of going into isolation. Close contacts of individuals who tested positive (particularly those co-housed with them) were also entered in the NU QI program. They were instructed to quarantine and were eligible to enroll in this study for up to 5 days after their last known exposure to an infected individual. All participants were also required to have received a negative nasal swab LAMP assay result 7 days prior to enrollment.

Individuals were recruited via either a link shared in an automated text message providing isolation information sent within 30 minutes of a positive test result, a call from a study recruiter, or a link shared by an enrolled study participant or included in information provided to all quarantining close contacts. In addition, signs/flyers were used at each testing location and a website was available to inform the community about the study.

Participants were required to be at least 18 years of age, have a valid university ID, speak English, have internet access, and live within 8 miles of the university campus. After enrollment and consent, participants completed an initial survey to collect information on demographics, vaccination status, prior infection history, and health history and were provided with sample collection supplies. Participants who tested positive prior to enrollment or during quarantine were followed for up to 14 days. Quarantining participants who continued to test negative by saliva RTqPCR (UIUC) or nasal swab RTqPCR (NU) were followed for up to 7 days after their last exposure. All participants’ data and survey responses were collected in the Eureka digital study platform.

### Sample collection

Each day, participants were remotely observed by trained study staff collecting:

1. 2 mL of saliva into a 50mL conical tube (UIUC study site only).
2. 1 nasal swab from a single nostril using a foam-tipped swab that was placed within a dry collection tube.
3. 1 nasal swab from the other nostril using a flocked swab that was subsequently placed in a collection vial containing viral transport media (VTM).

The order of nostrils (left vs. right) used for the two different swabs was randomized. For nasal swabs, participants were instructed to insert the soft tip of the swab at least 1 cm into the indicated nostril until they encountered mild resistance, rotate the swab around the nostril 5 times, leaving it in place for 10-15 seconds. After daily sample collection, participants completed a symptom survey. A courier collected all participant samples within 1 hour of collection using a no-contact pickup protocol designed to minimize courier exposure to infected participants.

### Saliva RTqPCR

After collection, saliva samples were stored at room temperature and RTqPCR was run within 12 hours of initial collection. The protocol for the covidSHIELD direct saliva-to-RTqPCR assay used has been detailed previously^14,15^. In brief, saliva samples were heated at 95°C for 30 minutes, followed by the addition of 2X Tris/Borate/EDTA buffer (TBE) at a 1:1 ratio (final concentration 1X TBE) and Tween-20 to a final concentration of 0.5%. Samples were assayed using the Thermo TaqPath COVID-19 Combo kit assay.

### Antigen testing

Foam-tipped nasal swabs were placed in collection tubes, transported with cold packs, and stored at 4°C overnight based on guidance from the manufacturer. The morning after collection, swabs were run through the Sofia SARS antigen FIA on Sofia 2 devices according to the manufacturer’s protocol.

### Nasal swab RTqPCR

For UIUC cohort, collection tubes containing VTM and flocked nasal swabs were stored at -80°C after collection and were subsequently shipped to Johns Hopkins University for RTqPCR and virus culture testing. After thawing, VTM was aliquoted for RTqPCR and infectivity assays. One ml of VTM from the nasal swab was assayed on the Abbott Alinity per manufacturer’s instructions in a College of American Pathologist and CLIA-certified laboratory. Calibration curve for Alinity assay was determined using digital droplet PCR (ddPCR) as previously described^16^.

### Virus culture from nasal swabs

Vero-TMPRSS2 cells were grown in complete medium (CM) consisting of DMEM with 10% fetal bovine serum (Gibco), 1 mM glutamine (Invitrogen), 1 mM sodium pyruvate (Invitrogen), 100 U/ml of penicillin (Invitrogen), and 100 μg/ml of streptomycin (Invitrogen)^17^. Viral infectivity was assessed on Vero-TMPRSS2 cells as previously described using infection media (IM; identical to CM except the FBS is reduced to 2.5%)^11^. When a cytopathic effect was visible in >50% of cells in a given well, the supernatant was harvested. The presence of SARS-CoV-2 was confirmed through RTqPCR as described previously by extracting RNA from the cell culture supernatant using the Qiagen viral RNA isolation kit and performing RTqPCR using the N1 and N2 SARS-CoV-2-specific primers and probes in addition to primers and probes for human RNaseP gene using synthetic RNA target sequences to establish a standard curve^18^.

### Viral genome sequencing and analysis

Viral RNA was extracted from 140 uL of heat inactivated (30 minutes at 95°C, as part of protocol detailed in^15^) saliva samples using the QIAamp viral RNA mini kit (QIAGEN). 100ng of viral RNA was used to generate cDNA using the SuperScript IV first strand synthesis kit (Invitrogen). Viral cDNA was then used to generate sequencing libraries using the Swift SNAP Amplicon SARS CoV2 kit with additional coverage panel and unique dual indexing (Swift Biosciences) which were sequenced on an Illumina Novaseq SP lane. Data were run through the nf-core/viralrecon workflow (https://nf-co.re/viralrecon/1.1.0), using the Wuhan-Hu-1 reference genome (NCBI accession NC_045512.2). Swift v2 primer sequences were trimmed prior to variant analysis from iVar version 1.3.1 (https://genomebiology.biomedcentral.com/articles/10.1186/s13059-018-1618-7) retaining all calls with a minimum allele frequency of 0.01 and higher. Viral lineages were called using the Pangolin tool (https://github.com/cov-lineages/pangolin) version 2.4.2, pango version 1.2.6, and the 5/19/21 version of the pangoLEARN model.

## Data Availability

All raw data will be made available at the time of publication in a peer-reviewed journal.

## Acknowledgments

We wish to thank Shumon Ahmed, Carly Bell, Nate Bouton, Shannon Bradley, Callie Brennen, Justin Brown, Coleco Buie, Emmaline Cler, Gary Cole, Trey Coleman, Alastair Dunnett, Darci Edmonson, Lauren Engels, Savannah Feher, Kelsey Fox, Lexi Freeman, Stacy Gloss, Yesenia Gonzalez, Montez Harris, Darcy Henness, Dan Hiser, Ayeshah Hussain, Daryl Jackson, Junko Jarrett, Michael Jenkins, Kalombo Kalonji, Syntyche Kanku, Steven Krauklis, Mary Krouse, Elmore Leshoure, Joe Lewis, Maggie Li, Angel Lopez, Guadalupe Lopez, Emily Luna, Chun Huai Luo, Colby Mackey, Skyler McLain, Yared Berhanu Melesse, Madison O’Donnell, Savanna Pflugmacher, Denver Piatt, Skyler Pierce, Jessica Quicksall, Gina Quitanilla, Crystal Reinhart, Ameera Samad, MacKenzie Scroggins, Monique Settles, Macie Sinn, Pete Varney, Evette Vlach, Raeshun Williams-Chatman, Jagadeesh Yedetore, and Todd Young for their efforts supporting recruitment, enrollment, logistics, and/or sample collection and processing at the UIUC study site.

We also wish to thank Kate Klein, Amelia Kelly, Nour Sayegh, Michael Govern, Mackenzie Furnari-Stickney, Sara Caudillo, Elizabeth Christain, Kristen Weber, James Jackson, Sowmya Ravi, Jack Novotny, Reno Stephens, Benjamin Engebretson, Patricia Helbin, Jessica Orozco, Alema Jackson, Adriana Quintatna, Hiba Abbas, Rana Saber, Eric Fickes, Matthew Butzler, Rocio Avila, Abhishek Agarwal, Robin Kim, William Zeng, Tosin Odumuye, Kirsten Knapton, Melissa Mongrella, Michael Newcomb, Kenyetta Sims, and Carol Govern for their efforts supporting recruitment, enrollment, logistics, and/or sample collection and processing at the NU study site.

We also thank Jeffrey Olgin, Noah Peyser, and Xochitl Butcher for assistance with the Eureka platform, Michelle Lore for assistance with REDcap, Melanie Loots for assistance with administration, Gillian Snyder for assistance in development of study protocols and logistics, and Erin Iturriaga and Jue Chen for study protocol development. Finally, we are grateful to Dr. Alvaro Hernandez and Ms. Chris Wright of the DNA Services Lab within the Roy J. Carver Biotechnology Center for expert assistance in establishment of a SARS-CoV-2 genomic sequencing protocol. Vero-TMPRSS2 cells were kindly provided by the National Institute of Infectious Diseases, Japan. Sofia 2 devices and associated supplies were provided to Carle Foundation Hospital by Quidel, however Quidel played no role in the design of the study or the interpretation or presentation of the data.

