## Supplemental Table 1 for "Longitudinal analysis of SARS-CoV-2 vaccine breakthrough infections reveal limited infectious virus shedding and restricted tissue distribution"

**Table S1: Study cohort details.**

| Participant ID: | Study site: | Viral Lineage: | Vaccination status: |
|---|---|---|---|
| 461913 | UIUC | B.1.596 | Moderna/mRNA-1273; **Within 14 days of first dose** |
| 467420 | UIUC | N.D. | Pfizer/BioNTech/BNT162b2; **Partially vaccinated** |
| 471645 | UIUC | N.D. | Moderna/mRNA-1273; **Within 14 days of first dose** |
| 471876 | UIUC | B.1.1.7 | Pfizer/BioNTech/BNT162b2; **Partially vaccinated** |
| 475670 | UIUC | P.1 | Pfizer/BioNTech/BNT162b2; **Fully vaccinated** |
| 481242 | UIUC | P.1 | JNJ-78436735; **Within 14 days of first dose** |
| 481672 | UIUC | N.D. | Pfizer/BioNTech/BNT162b2; **Within 14 days of first dose** |
| 482828 | UIUC | B.1.1.7 | Moderna/mRNA-1273; **Within 14 days of first dose** |
| 484249 | UIUC | B.1.1.7 | Pfizer/BioNTech/BNT162b2; **Partially vaccinated** |
| 485880 | UIUC | N.D. | JNJ-78436735; **Fully vaccinated** |
| 485948 | UIUC | N.D. | Moderna/mRNA-1273; **Within 14 days of first dose** |
| 486422 | UIUC | B.1.1.7 | JNJ-78436735; **Within 14 days of first dose** |
| 487250 | UIUC | B.1.1.7 | Moderna/mRNA-1273; **Fully vaccinated** |
| 487941 | UIUC | N.D. | Moderna/mRNA-1273; **Fully vaccinated** |
| 480838 | NU | B.1.623 | Pfizer/BioNTech/BNT162b2; **Within 14 days of first dose** |
| 481464 | NU | P.1 | Pfizer/BioNTech/BNT162b2; **Within 14 days of first dose** |
| 481706 | NU | N.D. | Pfizer/BioNTech/BNT162b2; **Fully vaccinated** |
| 487267 | NU | B.1.1.7 | Pfizer/BioNTech/BNT162b2; **Within 14 days of first dose** |
| 487402 | NU | B.1.1.7 | Pfizer/BioNTech/BNT162b2; **Within 14 days of first dose** |
| 487765 | NU | B.1.1.7 | Moderna/mRNA-1273**; Partially vaccinated** |
| 487958 | NU | N.D. | Pfizer/BioNTech/BNT162b2**; Partially vaccinated** |
| 488414 | NU | B.1.2 | JNJ-78436735**; Fully vaccinated** |
| 488687 | NU | B.1.1.7 | Pfizer/BioNTech/BNT162b2**; Partially vaccinated** |

**N.D. = Not determined**
